# Individual adherence to mass drug administration in neglected tropical disease control: a probability model conditional on past behaviour

**DOI:** 10.1101/2020.04.17.20069476

**Authors:** Robert J. Hardwick, James E. Truscott, William E. Oswald, Marleen Werkman, Katherine E. Halliday, Rachel L. Pullan, Roy M. Anderson

## Abstract

We present a general framework which describes the systematic (binary) scenario of individuals either taking treatment or not for any reason, over the course of mass drug administration (MDA) — which we refer to as ‘adherence’ and ‘non-adherence’. The probability models developed can be informed by observed adherence behaviour as well as employed to explore how different patterns influence the impact of MDA programmes, by the use of mathematical models of transmission and control. We demonstrate the interpretative value of the developed probability model employing a dataset collected in the TUMIKIA project, a randomised trial of deworming strategies to control soil-transmitted helminths (STH) by MDA conducted in coastal Kenya. We stratify our analysis by age and sex, although the framework which we introduce here may be readily adapted to accommodate other stratifications. Our findings include the detection of specific patterns of non-adherence in all age groups to varying extents. This is particularly apparent in men of ages 30+. We then demonstrate the use of the probability model in stochastic individual-based simulations by running two example forecasts for the elimination of STH transmission employing MDA within the TUMIKIA trial setting with different adherence patterns. This suggested a substantial reduction in the probability of elimination (between 23-43%) when comparing observed adherence patterns with an assumption of independence, with important implications for programmes. The results here demonstrate the considerable impact and utility of considering non-adherence on the success of MDA programmes to control neglected tropical diseases (NTDs).

**Author summary:** Mass drug administration (MDA) is an important tool in the prevention of morbidity caused by various NTDs and in the reduction of their transmission. Due to a variety of social and behavioural reasons, many people will either not be offered or refuse such treatment, and if this behaviour is recurring at an individual level, then control measures may face a challenge in achieving their stated goals. Accurately describing the patterns of individual adherence or non-adherence to MDA control measures for NTDs from data, followed by their use in simulated scenarios is a relatively recent development in the study of NTDs. Past analyses assessing individual adherence have informed the approach we take in this work. However, we have sought to provide a framework which encapsulates as many types of adherence behaviour as possible to facilitate the assessment of impact in mathematical models of parasite transmission and control. Our example application to the TUMIKIA data highlights the importance of such a general framework as we find a dependence on past behaviour that may have been missed in standard statistical analyses.

## 1 Introduction

Recent reviews, guidelines and analyses predicting the outcome of mass drug administration (MDA) to control the transmission of various neglected tropical diseases (NTDs) all emphasise the importance of individual adherence in successfully reaching elimination targets [1–9]. Such analyses have taken a variety of approaches in describing how participants in a given MDA programme with multiple rounds can either not be offered, or actively avoid, treatment in a potentially repetitive manner. There are a wide range of published studies of treatment adherence in the literature and mathematical plus statistical models of adherence are included in micro-simulations of infectious agent disease control strategies across a great variety of infectious agents including HIV, tuberculosis and NTDs [1, 3–8, 10]. A much larger literature exists for non-infectious diseases such as blood pressure control and statin use.

Although a range of terms have been used to describe this phenomenon [7], here we refer to the binary scenario of individuals either taking treatment or not, for any reason, over the course of multiple rounds of MDA as ‘adherence’ or ‘non-adherence’. Ultimately, the effect that this behaviour has on the success or failure of control through MDA is of great importance and not fully recognized in policy formulation concerning the monitoring and evaluation of MDA programmes by WHO and national governments.

In this paper, we develop a general approach to describe individual adherence or non-adherence to MDA. Our principal aim is to provide a framework within which as many patterns or adherence behaviour as possible are captured by a general probability model so that the evaluation of the importance of adherence patterns on the impact of MDA programmes can be precisely quantified. This also involves employing, for example, models of parasite transmission and control by MDA. To illustrate how our methodology may be implemented and interpreted in practice, we apply it to data collected during the TUMIKIA project: a recent cluster randomised, controlled trial of the impact of MDA on the transmission of STH infections in Kwale County, Kenya [11–13]. A statistical analysis has already been performed on this dataset, as described in Ref [13], and so the analysis we present here is to illustrate the application of our probability model only.

## 2 Methods

### 2.1 Model definitions

In this section, we will lay out a general probability model for the treatment of adherence across multiple rounds in an MDA intervention programme. In Appendix S3, we discuss other implementations of adherence in probability models and how they fit within our general framework.

At the level of an individual involved in an MDA treatment programme, we describe adherence as a binary ‘choice’, made at each round of MDA, of whether to receive treatment or not (or if treatment is, or is not, accessible to the individual). We associate a probability with this ‘choice’, which is composed of both an individual’s access to treatment and their personal choice to take it, making each round a Bernoulli trial for each individual.

We identify three main ways in which the probability of adherence can vary in a population over the course of an MDA intervention.

1. Dependence on past behaviour: An individual’s probability of adhering in the current round may depend on their individual history of adherence in past rounds. This could be alternating (for example, being treated in the previous round may make individuals feel their participation in this round is less important, or prior experience of unpleasant side-effects may prevent adherence) or may be persistent (for example, those who live in hard-to-reach or marginalised households may be consistently missed).
2. Time dependence: everyone involved in the trial may be subject to external influences that change over time. For example, enthusiasm or funding for the treatment programme may decline as it proceeds, or unforeseen sociological or political events may change the population’s inclination to take part in the programme. This will result in the probability of treatment in a given round for a given individual being explicitly dependent on time and is distinct from dependence on past behaviour.
3. Population-level heterogeneity: the probability of adherence may vary systematically across the population on the basis of socio-demographic stratifying characteristics. That is, individuals may have a individual probability of adherence that they retain across multiple rounds of the intervention. In this case, the probability of adherence will have a distribution across the population. Typically, population-level heterogeneity may be strongly correlated with covariates such as sex or age, in which case it can be represented by a stratification of the population into sub-groups, each with their own adherence probability.

In reality, any model of adherence might include one or more of these sources of variability, or none at all in the default case in which the adherence probability is constant across all individuals and all treatment rounds and does not depend on past history. For the purpose of illustration, we can create a tree of possible model types based on the possible sources of variability (see Fig 1). Models can be stratified into types which have some degree of dependence on the past behaviour of individuals and those that do not. Within each group, there are models with and without population heterogeneity in adherence (heterogeneous and homogeneous populations, respectively) and those with and without time-dependent adherence probabilities (time-dependent and independent, respectively).

**Fig 1.**
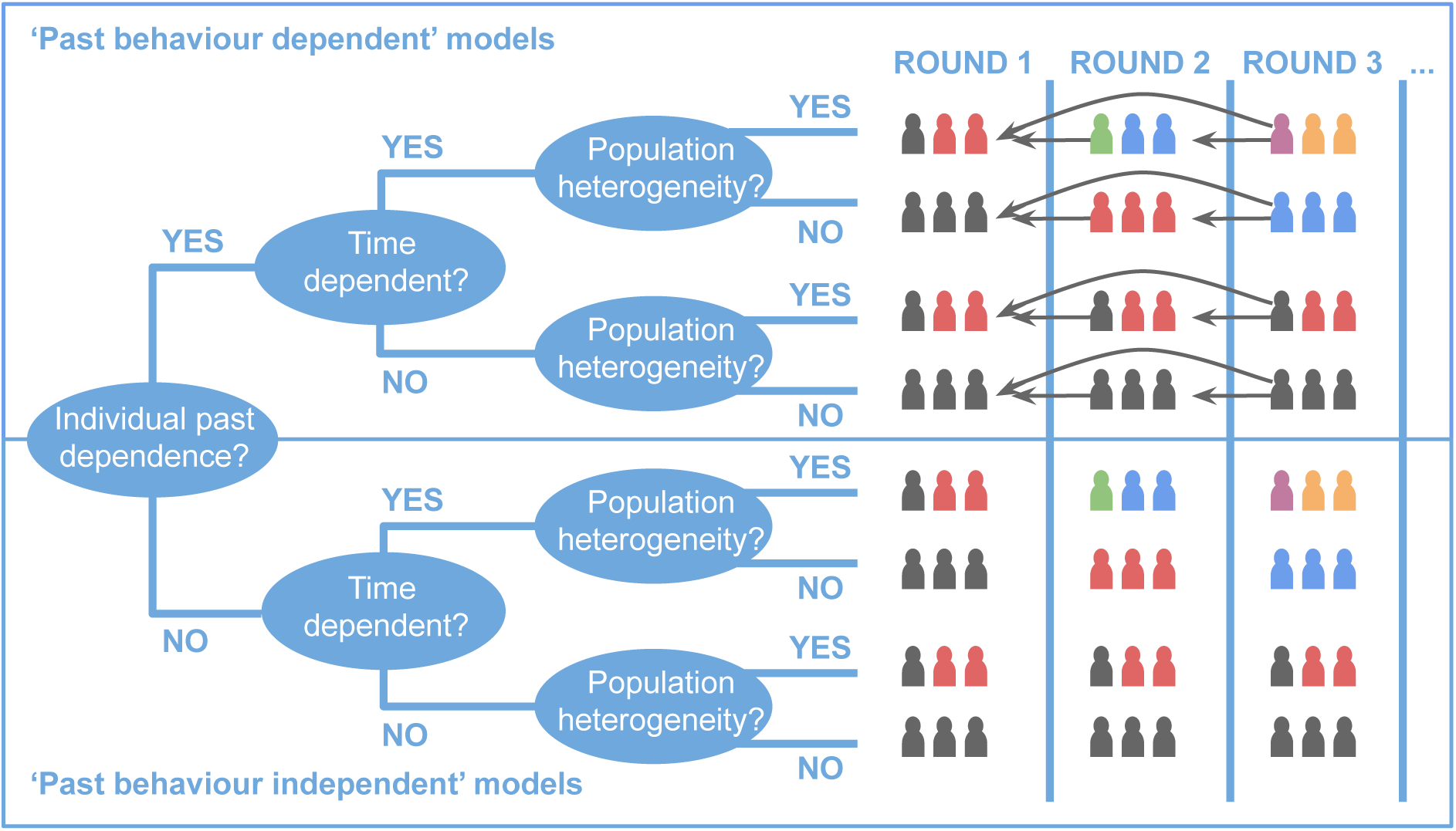
A decision tree illustrating the possible classes of behaviour which may be characterised in adherence models. In the illustrations, colours of individuals denote unique probabilities of receiving treatment and arrows to previous rounds denote dependency of these probabilities on past behaviour.

The distinctions above are of critical importance as it is possible for example for a treatment programme to suffer severely from past behaviour-dependent non-adherence without any apparent heterogeneity in adherence within the population. They also allow us to categorise and clarify models of adherence already described in the literature [1, 4–8].

The Plaisier model assigns a probability of adherence to each individual which they then retain for the duration of the MDA programme [14, 15]. As such, this model would be characterised by us as a heterogeneous population, time-independent model with no explicit individual dependence on past behaviour. We discuss the relationship of the Plaisier model (and others [5]) to our categorisation of adherence models with more detail in Appendix S3. Additional technical details and calculations may also be found in Appendix S1 for the three adherence categories.

### 2.2 Individual past behaviour-dependent adherence

#### 2.2.1 Basic model

When the probability of an individual taking treatment is not dependent upon any of their past behaviour, then it is simply given by the coverage *c*_*n*_ in each round *n* of MDA. In the absence of population heterogeneity, this probability would then apply to all individuals within a given cohort — a case which corresponds to either the 6th or 8th row of Fig 1, depending on whether the coverage changes over time, i.e., between rounds.

For the case of a homogeneous population with a dependence on past behaviour between successive rounds, let us consider the dynamics of a single individual. In this case, the model becomes a simple Markov chain. The possible patterns of adherence behaviour by an individual after two successive rounds of treatment are TT, TF, FT and FF, where T and F are receiving and not receiving treatment, respectively. Let the probability of receiving treatment in the first round be set to *P* (T) = *α*. In round 2, we now fix the conditional probability of getting treated, given treatment in the first round as *P* (T′| T) = *β*. Let us also set a corresponding conditional probability for not being treated in the second round given that there was no treatment in the first round as *P* (F′|F) = *γ*. As such, *β* and *γ* are now measures of consistent behaviour.

To avoid unnecessary repetition, we shall once again use the notation *p*_*n*_ to denote the probability of treatment in the *n*-th round. Assuming that the conditional probabilities *β* and *γ* are constant, the time-independent Markov model may be mapped to the following recursion relation

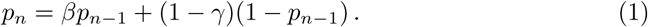

In Appendix S1, we demonstrate how to obtain the following solution to Eq (1)

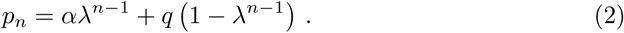

Where we have defined

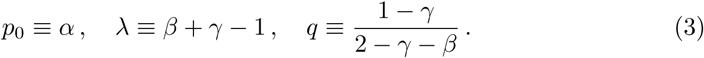

Notice that by matching *p*_*n*_ to the coverage of treatment in a given population, one may directly compare the impact of adherence models such as Eq (1) to those with past behaviour-independent adherence. Furthermore, by setting *λ* = 0 in Eq (2) one finds the model for past behaviour-independent adherence that is time-independent, i.e., *p*_*n*_ = *β* = 1 − *γ*.

Any sequence of treatments can be seen as a set of alternating adherent and non-adherent runs. A key statistic in the context of preventive chemotherapy is the run length (in rounds) over which an individual adheres or fails to adhere. For an adherence run, this is the number of consecutive treatment adherences, given an initial adherence. This can also be thought of as the first passage time to failure. Since the *P* (T′| T) = *β* is constant, the run length is distributed according to a geometric distribution, with

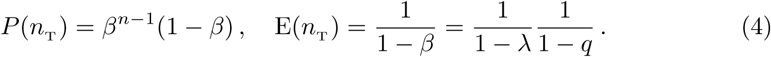

Correspondingly, for a run of failures,

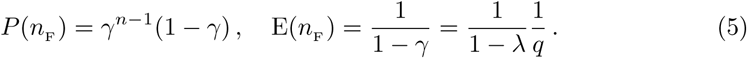

Any long run of treatment choices by an individual will breakdown into an alternating sequence of F and T runs. Hence, the probability of a round chosen at random being T, *P* (T), is

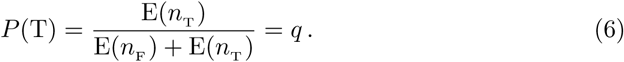

From Eqs (4) and (5), it is clear that as *λ* approaches 1, the length of both success and failure runs grows as 1*/*(1 − *λ*). In the absence of past behaviour dependence, *λ* = 0 and the adherent and non-adherent run lengths are given by 1*/*(1 − *q*) and 1*/q*, respectively.

#### 2.2.2 Statistical inference from data

Let us now only consider two rounds of treatment to illustrate how we may calculate the important quantities for statistical inference of the time-independent Markov model from a real dataset. Recall that the possible patterns of adherence behaviour by an individual after two successive rounds of treatment are TT, TF, FT and FF, where T and F are receiving and not receiving treatment, respectively. Once again, let: the probability of treatment in the first round be set to *P* (T) = *α*; the conditional probability of getting treated in the second round, given treatment in the first round be set to *P* (T′| T) = *β*; and the conditional probability for not being treated in the second round given that there was no treatment in the first round be set to *P* (F′|F) = *γ*.

In this model, there are effectively 4 types of people with probabilities and behaviours, mapped out in Table 1. Using the probability table, one may infer directly that the the likelihood *ℒ* (*D*|***θ***) of the data *D* = {*N*_T_, *N*_F_, *N*_TT_, *N*_TF_, *N*_FT_, *N*_FF_} — where *N*_T_ and *N*_F_ are the number treated and not treated in the first round and *N*_TT_ is the number treated in the first and the second rounds, etc. — is a multinomial distribution, where ***θ*** ∈ Ω_***θ***_ is now a 3-vector defined over the model parameter space ***θ*** = (*α, β, γ*) within the prior domain Ω_***θ***_ = {***θ*** | *θ*_1_ ∈ [0, 0], *θ*_2_ ∈ [0, 1], *θ*_3_ ∈ [0, 1]. The multinomial can then be factored into independent functions of the three parameters, such that

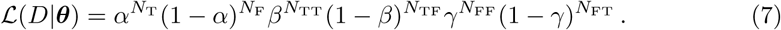

**Table 1.** Probability table corresponding to two successive rounds of treatment.

| Behaviour | Probability |
| --- | --- |
| TT | $\alpha\beta$ |
| TF | $\alpha(1 - \beta)$ |
| FT | $(1 - \alpha)(1 - \gamma)$ |
| FF | $(1 - \alpha)\gamma$ |

The likelihood above is effectively three independent beta distributions, one in each of the parameters, such that the posterior distribution *𝒫* (***θ***|*D*) becomes

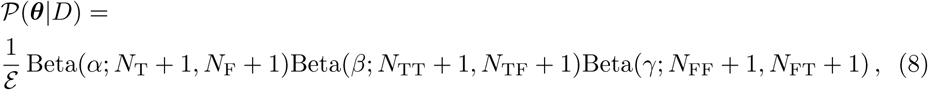

where we have assumed a flat prior *π*(***θ***) ∝ 1 to derive the following Bayesian evidence normalisation

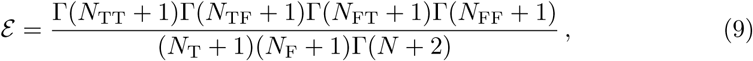

and *N* = *N*_T_ + *N*_F_ is defined as the total number of individuals.

Note here that Eqs (7) and (9) may be generalised to the case where *n* rounds of treatment have taken place. We have provided these expressions in Appendix S1.

### 2.3 Time-dependent adherence and more general behaviour

#### 2.3.1 Introducing the choice matrices

A significant generalisation of Eq (1) introduces the lower triangular matrices, with elements 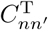 and 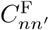 corresponding to the conditional probabilities of treatment and non-treatment in round *n* given treatment and non-treatment in round *n*′, respectively, such that

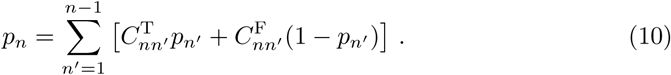

We shall hereafter refer to the above matrices as ‘choice matrices’. In Appendix S1 we demonstrate that the model parameterisation defined in Eq (10) is extremely general — encapsulating all of the possible adherence behaviours illustrated in Fig 1.

#### 2.3.2 Lower diagonal choice matrices: the time-dependent Markov model

When the only nonzero elements of the choice matrices in Eq (10) are along the lower diagonals, i.e., such that only 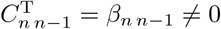 and 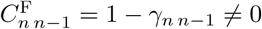, the system is described by a time-dependent Markov process with recursion relation

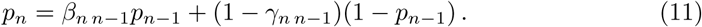

Following a similar argument to the one used in solving the homogeneous Markov model (which is provided in detail in Appendix S1), we may obtain a solution to Eq (11), which is given by

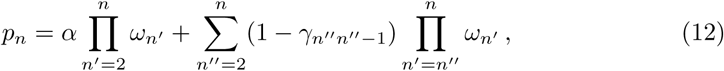

where we have defined an important new quantity

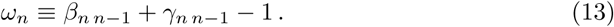

Notice, firstly, that when *ω*_*n*_ = 0 the system reverts to a time-dependent past behaviour-independent adherence model, i.e., without past behaviour dependence such that *p*_*n*_ = *β*_*n n*−1_ = 1 − *γ*_*n n*−1_. By analogy with the time-independent Markov model, |*ω*_*n*_| ≠ 0 signals the presence of some degree of past behaviour-dependent adherence behaviour. In more detail, for successive rounds over which *ω*_*n*_ > 0, the system will relax towards the steady state and when *ω*_*n*_ < 0 this will be accompanied by oscillatory behaviour. Note also that *ω*_*n*_ may act as an indicator for the severity of adherence and non-adherence behaviour in the system — where larger absolute values for *ω*_*n*_ approaching a maximum of 1 will indicate increasingly past behaviour dependence.

At the extrema of: *ω*_*n*_ = 1, individuals repeat their past behaviour exactly and indefinitely, i.e., TTTTT… and FFFFF…; and *ω*_*n*_ = −1, individuals repeat the opposite of their past behaviour exactly and indefinitely, i.e., TFTFT… and FTFTF. The value of *ω*_*n*_ is therefore a useful indicator for the type of adherence behaviour in the relatively general description of time-dependent Markov models.

#### 2.3.3 Fitting the model to adherence data

The universality of the choice matrix approach suggest that it is an ideal candidate for parameterisation of the inference problem from data and model comparison. Let the data now correspond to a set of *n*-vectors *D* = {***X***} where each individual’s adherence or non-adherence behaviour in the *n*-th round is recorded, such that *X*_*n*_ = T, F. Using Eq (10) the full generalisation of the likelihood (which supports all of the possible adherence models) becomes

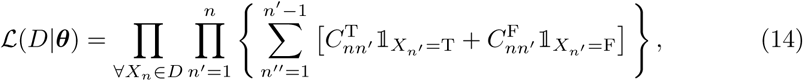

where 𝟙_*A*_ denotes an indicator function which takes value unity when condition *A* is satisfied, else it vanishes.

The large number of available degrees of freedom in Eq (14) motivates a systematic approach to inferring the choice matrix components from a given set of data. To isolate the many degrees of freedom and compared between the relative evidence for them in the data, we construct models where past behaviour-dependent adherence only occurs for a single round and is past behaviour dependent to only one other round. In the choice matrix, all other degrees of freedom are assumed to be time-dependent past behaviour-independent adherence, i.e. 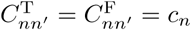. The likelihoods and Bayesian evidence normalisations for this more restricted set of models are calculated in Appendix S1.

### 2.4 Population heterogeneity in adherence

The probability of adherence may vary greatly across a population of individuals. The first possible form that this heterogeneity may take can be attributed to age, gender and social plus behavioural factors. In such cases, stratification of the population into separate cohorts for study is an appropriate tool to quantify this variation.

The second possible form that population heterogeneity could take may not be immediately attributable to social or demographic groupings. In such situations, the adherence probability for an individual can be drawn from a distribution which applies to the entire population or defined sub-group of the population within the study. This approach is the same as used in other models in the literature (see Appendix S3 for more details). We shall now briefly elaborate on how one might include this form of heterogeneity in the formalism we have introduced in this work through a simple, generic example.

To illustrate the generic effect of the population heterogeneity described above on our individual adherence probabilities, let us consider the time-independent Markov model we introduced earlier. The long-term probability of adherence *q* in Eq (2) may itself be randomly drawn from a population heterogeneity distribution *P*_pop_(*q*) for an individual within the specified cohort of study, such that *q* ∼ *P*_pop_(*q*). Note also that *λ* in Eq (2) need not vary between individuals at the same time. Using the results given in Eqs (4) and (5) for the same model one may deduce that the mean adherent and non-adherent run lengths are modified by

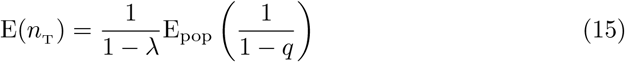

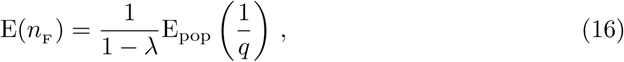

where E_pop_(·) denotes taking an expectation value with the distribution *P*_pop_(*q*). Hence, depending on the choice for this distribution, one may either shorten or lengthen the mean run lengths across the population accordingly. Note that due to the fact that *q* is a probability, a possible and widely applicable candidate for *P*_pop_(*q*) is the beta distribution.

### 2.5 Dataset used

In the four rounds of individual adherence data recorded in the TUMIKIA project of MDA to control STH infections, we have split the study population into pre-school-aged children (pre-SAC, ages 0-4 — where only ages 2-4 were eligible for treatment), school-aged children (SAC, ages 5-14) and other adult age categories. The individuals considered in the TUMIKIA dataset comprise a randomly sampled cohort of 21978 individuals living in 40 ‘community units’ (each constituting around 1000 households) in the biannual treatment arm, in which albendazole was targeted to all individuals aged over two years during house-to-house delivery campaigns conducted by community health volunteers every six months. Importantly, the movement of individuals between age groups is not explicitly specified in the model, and so some time variation in the inferred compliance behaviour may be attributable to the process of an individual moving into a new age category over the course of the trial.

## 3 Results

### 3.1 Overview of statistical analysis

In Fig 2 we provide one example set of plots from Appendix S2 for the SAC (4-15) age group of the TUMIKIA adherence dataset which gives the maximum likelihood as well as the limits of the marginalised 95% credible region for the conditional probabilities given treatment (filled points) or non-treatment (hollow points) in a previous round of the overall, male and female participants in the top, middle and bottom rows, respectively. Note that we are using the symbolic representation for behaviours which we introduced in Sec 2 — receiving treatment in a given round is denoted by a ‘T’, whereas not receiving treatment in a given round is denoted by an ‘F’. In the left column the constant conditional probabilities between any given sequential pair of rounds have been inferred, which corresponds to the time-independent probability model. In the right column all possible round pair dependencies are considered (indicated by the arrows on the horizontal axis), where in each case the components corresponding to a given round were measured assuming all other respective rounds were inferred to be from past behaviour-independent adherence. In all plots, above each pair of components we have also provided the log-Bayes factors [16] (see Appendix S2 for further explanation), where the evidence for has been evaluated using the relations provided in Appendix S1 and the reference model evidence has been set to that of time-dependent past behaviour-independent adherence for all components.

**Fig 2.**
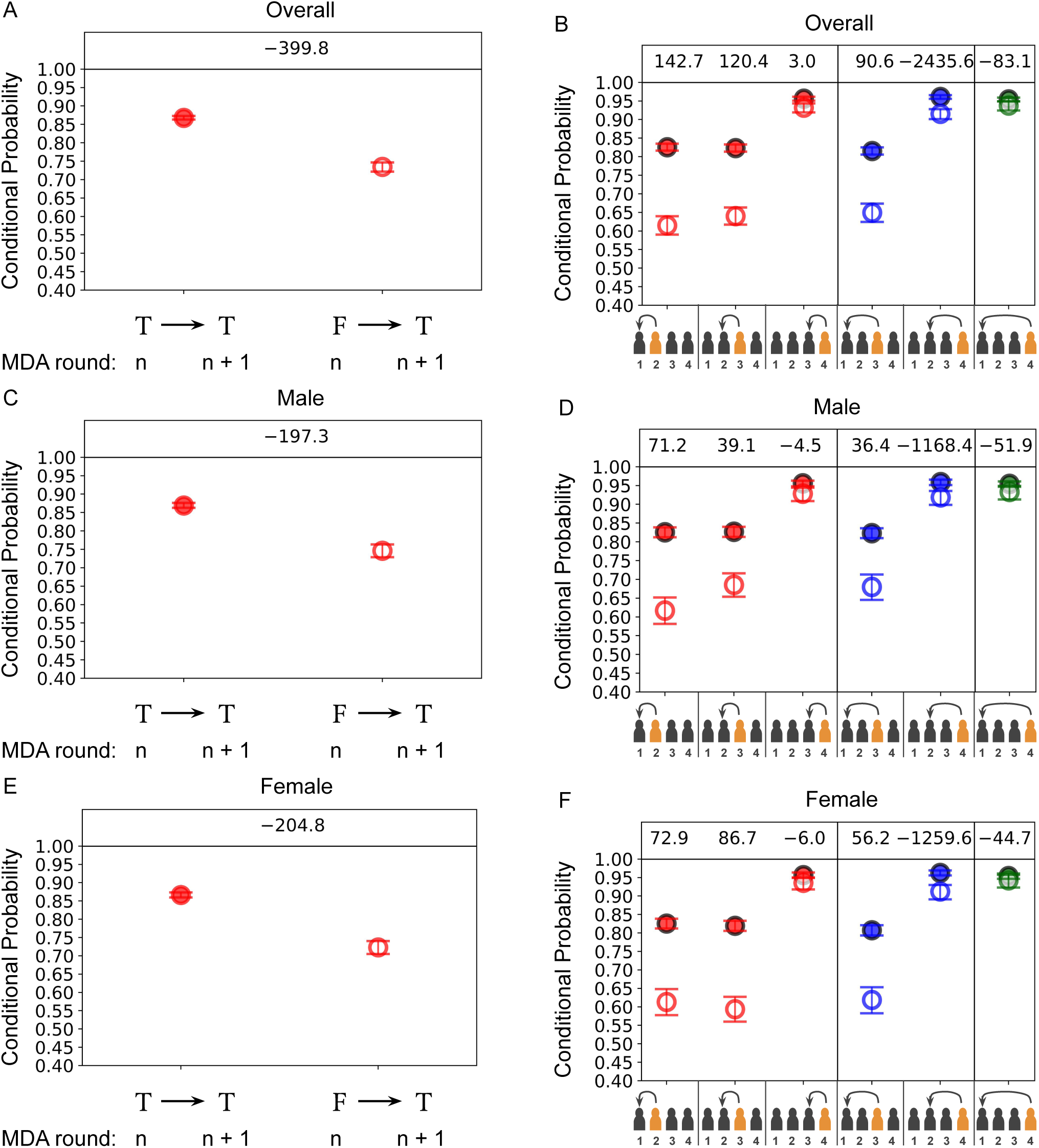
*Left column:* The maximum likelihood as well as the limits of the marginalised 95% credible region for the conditional probabilities of receiving treatment for any given pair of sequential rounds (these are hence homogeneous in time and the process is Markovian) given treatment (filled points) or non-treatment (hollow points) in a previous round. *Right column:* The same as the left column but with allowed time-dependent in the conditional probabilities of receiving treatment in each respective round (highlighted in orange on the horizontal axes). In each case the components corresponding to a given round were measured assuming all other respective rounds were inferred to be from time-dependent past behaviour-independent adherence and hence the likelihood is given in Appendix S1. Different colours for each point correspond to different lengths in time for the dependencies in behaviour. The datasets used are from the standard SAC (4-15) age category from a cohort of individuals from the biannual treatment arm of the TUMIKIA project where the: top row corresponds to the overall group; middle row corresponds to the male sub-group; and bottom row corresponds to the female sub-group.

From Fig 2, the pre-SAC age group appears to be well-described by a time-dependent probability model and past behaviour-dependent non-adherence is clearly present. This may be identified by the largest log-Bayes factor values being given in the red-coloured right column plots for all three sets of plots. However, the conditional probabilities in all groups appear to drift closer together by round 4 of treatment, which signals a gradual transition from past behaviour-dependent to independent adherence. From these plots we also report no evidence for the existence of dependencies between rounds in the pre-SAC age group that depart from a Markovian description (as can be inferred from the comparatively small log-Bayes factors for the blue and green conditional probabilities in the right column of all plots). We also have a detailed description of all of these findings and those for all of the other age categories in Appendix S2.

In Table 2 we have also provided the *ω*_*n*_ values, calculated using Eq (13), for each age group and sex inferred from the TUMIKIA project dataset. This value was shown in Sec 2 to be an indicator of how past behaviour influences the adherent and non-adherent behaviour of individuals in a future round of MDA treatment, with larger (positive or negative) values corresponding to a greater degree of dependence on past behaviour (individuals systematically non-adhering and adhering with more probable repetition, respectively) and smaller (positive or negative) values corresponding to an individual’s adherence pattern showing less of a dependence on their past behaviour.

**Table 2.** A measure of how past behaviour influences the adherent and non-adherent behaviour of individuals is in the *n*-th round of treatment, *ω*_*n*_ ≡ *β*_*n n*−1_ + *γ*_*n n*−1_ −1, which was introduced in Eq (13). This value is given for each age group and sex inferred from the TUMIKIA project dataset and is computed using the maximum likelihood values for the conditional probabilities. The uncertainties quoted with each value correspond to the standard deviation of *ω*_*n*_ in each case.

| Age group | $\omega_2$ (Male) | $\omega_3$ (Male) | $\omega_4$ (Male) | $\omega_2$ (Female) | $\omega_3$ (Female) | $\omega_4$ (Female) |
| --- | --- | --- | --- | --- | --- | --- |
| Pre-SAC | $0.294 \pm 0.012$ | $0.241 \pm 0.011$ | $0.051 \pm 0.006$ | $0.237 \pm 0.013$ | $0.250 \pm 0.012$ | $0.046 \pm 0.006$ |
| SAC | $0.209 \pm 0.006$ | $0.141 \pm 0.006$ | $0.027 \pm 0.003$ | $0.213 \pm 0.007$ | $0.226 \pm 0.007$ | $0.021 \pm 0.003$ |
| 15-29 | $0.228 \pm 0.010$ | $0.210 \pm 0.010$ | $0.111 \pm 0.008$ | $0.223 \pm 0.009$ | $0.182 \pm 0.009$ | $0.066 \pm 0.007$ |
| 30-49 | $0.259 \pm 0.009$ | $0.308 \pm 0.009$ | $0.268 \pm 0.008$ | $0.223 \pm 0.009$ | $0.174 \pm 0.008$ | $0.118 \pm 0.007$ |
| 50+ | $0.244 \pm 0.010$ | $0.286 \pm 0.010$ | $0.259 \pm 0.009$ | $0.195 \pm 0.011$ | $0.181 \pm 0.011$ | $0.139 \pm 0.009$ |

We can see quite clearly from Table 2 by the values of the conditional probabilities that a degree of past behaviour-dependent non-adherence is indeed present in all the age groups, with the exception of the final round *ω*_4_ values for those in the pre-SAC (which have mostly aged into SAC by this point) and SAC categories. This effect is explained in more detail by Ref [13]. Table 2 also shows that the most past behaviour-dependent non-adherent age group and sex appears to be males aged 30+ (they have the largest conditional probability values across all rounds of treatment). In addition to these results, Eq (1) appears to require extension to an equivalent time-dependent model — see Eq (11) — in order provide a good descriptive model for many of the past behaviour-dependent non-adherent age groups and sexes.

### 3.2 The impact of adherence on forecasts

In this section we illustrate the impact of adherence, as described by our probability model, on the predictions made by simulations for the impact of MDA on the chances of elimination of transmission by considering the case study of TUMIKIA and forward-projecting the impact of continued MDA treatment. The adherence models fitted to the TUMIKIA adherence data in the previous section are applied within a stochastic individual-based simulation of hookworm transmission for two example communities that were treated in the TUMIKIA project [17]. The resulting effect that the known TUMIKIA adherence has on the transmission elimination probability of hookworm in these two clusters is given in Table 3, where an equivalent transmission elimination probability assuming past behaviour-independent adherence is also provided for direct comparison in each case. The results we show here suggest a substantial reduction in the probability of elimination (between 23-43%) when comparing observed adherence patterns with an assumption of independence. This finding has important implications for MDA programmes as it is clear for progress to be made (and indeed quantified) towards transmission elimination in communities, an additional priority must be placed in accurately discerning the local patterns of adherence.

**Table 3.** The transmission elimination probability evaluated by fully age-structured stochastic individual-based simulations of hookworm (with adult worm and eggs/larvae mortality rates set to *μ*_1_ = 0.5 and *μ*_2_ = 26.0 per year, respectively and the density dependent fecundity factor is set to *γ* = 0.01, as considered in Ref [17]) with two different clustered community types specified by the TUMIKIA transmission parameters inferred from the baseline epidemiological data in Ref [17]. The parameters quoted are the endemic prevalence *P*, parasite aggregation parameter *k*, basic reproduction number *R*_0_ and cluster population number *N*, where the age profiles are all assumed to be exactly flat for simplicity. The transmission elimination probabilities are evaluated after 100 years post-cessation of MDA and are quoted assuming either past behaviour-independent adherence (i.e., simple time-dependent coverage in age groups and only population heterogeneity at the level of age bins) or the adherence behaviour inferred from our model in this paper for the TUMIKIA project (see Appendix S2). In parameter set 1: (*P, k, R*_0_, *N*) = (0.15, 0.05, 2.1, 1000) and parameter set 2: (*P, k, R*_0_, *N*) = (0.4, 0.15, 2.5, 1000).

| Community type (see Ref [17]) | Transmission elimination probability<br>(Past independent adherence) | Transmission elimination probability<br>(TUMIKIA adherence) |
| --- | --- | --- |
| 1: Lower baseline prevalence, less intense transmission but higher aggregation. | 0.582 | 0.148 |
| 2: Higher baseline prevalence, more intense transmission but lower aggregation. | 0.902 | 0.672 |

Note that the TUMIKIA adherence pattern that we have used in these representative clustered communities has been inferred across all clusters. For a specific forecast of the TUMIKIA trial outcome, one should input both cluster-level posterior uncertainties on the deworming simulation parameters at baseline as well as the cluster-level inferred adherence model parameters — from which one might expect even more substantial heterogeneity in outcomes at the cluster level. The impacts we quote here are representative of the significance of adherence patterns to programs, rather that specific forecasts for the TUMIKIA trial clusters.

## 4 Discussion and conclusions

The causes for non-adherent behaviour are undoubtedly varied as a result of both the type of treatment, and social plus behavioural factors in any defined treated community. In this paper we have been able to develop a simple but comprehensive framework which describes the systematic binary ‘choice’ of individuals to either take treatment, or not for any reason, over the course of multiple rounds of MDA — which we have referred to as ‘adherence’ and ‘non-adherence’, respectively.

Here, we introduce a flexible adherence framework, which can be used to account for a range of behaviours not yet considered in existing models of MDA. This analysis defines a summary parameter, *ω*_*n*_, which can be used as a guide to indicate the strength of adherent or non-adherent behaviour in any given sitting. An equivalent, frequentist interpretation for this parameter is that of a correlation coefficient between the (binary) behaviour of an individual being treated (1) or not (0) in the *n*-th round of MDA and their behaviour in the (*n* − 1)-th round of MDA.

In order to demonstrate the application of our probability model, we applied it to the recently collected adherence data from the TUMIKIA project in Kenya, which aims to control STH infections by repeated drug treatment, in Sec 3. Findings from the analyses presented here extend and support previous work [13], which include past behaviour-independent adherence or non-adherence for school-aged children (SAC) and the detection of past behaviour-dependent non-adherence to treatment in nearly all other age groups and both sexes. A full description of our results and analysis is given in Appendix S2.

The validity of interpreting the inferences made in Sec 3 as directly due to individual behaviour patterns using the TUMIKIA project adherence data [13] should be considered carefully. An important caveat to this interpretation is that, for various reasons, some individuals were not offered treatment and were hence automatically accounted for as ‘non-adherent’ within the data, and as such we cannot discriminate between refusing, and not being offered, treatment. The impact of these two reasons for non-adherence to the success of an MDA programme is however the same, and hence, the practical use of inferring this pattern of adherence for simulation forecasts of MDA outcome is still appropriate. We cannot discriminate this behaviour pattern from simply not being offered treatment in the TUMIKIA dataset.

Using the adherence behaviour patterns for a series of age groups from the Kenyan TUMIKIA dataset, we then demonstrated the use of a stochastic individual-based simulation model for STH transmission and control by MDA by running two example forecasts for the likelihood of elimination of hookworm transmission with the adherence behaviour recorded in Kenya by comparison with runs that assume random adherence at each round of treatment for any given treatment coverage level. From Table 3 it is immediately clear that although there is relatively high coverage of MDA in the TUMIKIA project [11, 12], the occurrence of past behaviour-dependent non-adherence has an important effect on the transmission elimination probabilities, shifting the chances of hookworm elimination in both communities lower by 43% and 23%, respectively, when compared to the standard forecasts which assume that current and future adherence behaviour is independent of past adherence. The difference between the two assumptions is striking. These results show clearly the great importance of measuring adherence if the outcome of any given MDA based control programme is to be correctly predicted. Note that a full forecast of TUMIKIA would also include posterior uncertainties evaluated for the simulation parameters and initial conditions, hence the simulation results we have provided in this work are intended only as guide to the importance of including adherence patterns into forecasting simulations.

In Table 3 when the baseline (equilibrium) prevalence is lowered, but the aggregation increases, this typically has the seemingly counter-intuitive effect of lowering the overall probability of eliminating transmission. This result arises from the relationship we have assumed between baseline (equilibrium) prevalence and the aggregation of worms within hosts, which was statistically inferred from the TUMIKIA data in Ref [17]. When aggregation of worms within hosts is higher for the population at equilibrium, this means that fewer people are required to sustain transmission indefinitely. In contrast, when the aggregation of worms within hosts is lower for the population at equilibrium, this means that a larger number of individuals are required to sustain transmission indefinitely.

For perspective on how our new adherence framework fits into the wider context of analysis approaches, we investigated the Plaisier model, which assigns a probability of adherence to each individual which they then retain for the duration of the MDA programme [14, 15]. In our categorisation scheme — which is illustrated in Fig 1 — this model would be characterised by us as a heterogeneous population, time-independent model with no explicit individual dependence on past behaviour. We also discussed the relationship of other models [5] to our categorisation of adherence models, where all of our comparisons can be found in Appendix S3.

WHO recommendations on how best to measure the impact of MDA programmes to control NTDs only advise recording patterns of treatment coverage round by round with some rough stratification by the age groupings treated (usually pre-SAC, SAC and adults). No advice to Ministries of Health is given on trying to record adherence patterns in part because of the challenges presented in recording these patterns in many communities. As we have demonstrated, the precise form of the adherence pattern can greatly influence the extent of the required MDA coverage and the number of treatment rounds necessary to eliminate parasite transmission. For instance, as we demonstrated with data collected from the TUMIKIA project in Sec 3.2, if the individual adherence patterns are found to have dependence on past behaviour, this can significantly reduce the probability of transmission elimination.

The lack of WHO guidance on adherence is understandable, given the costs and time involved in longitudinal studies to record adherence of individuals within any given MDA programme. However, given the importance of these pattens in determining control progamme impact and outcome, collecting such data should be given a higher priority even if just focused on a few sentinel sites to broadly capture the prevailing behaviours in defined settings. It is also likely, the social, environmental and other influences will create some heterogeneity in adherence patterns within countries and health implementation units. Additional background research on what degree of heterogeneity exists in a given country would also be of great value. In the coming few years more data on adherence patterns will emerge from detailed research studies of MDA impact to add to the information provided by the TUMIKIA study [11]. These include the ongoing DW3 trial studies in India, Benin and Malawi for the control of STH [18] and the Geshiyaro study in Ethiopia for the control of STH and schistosome infections by MDA [19].

Though we have addressed in this paper how those equipped with modelling capacities should use this data to improve forecasting pipelines, it remains an open, and important, question as to how programme implementers can best use the data directly. We propose in this case that computing the binomial frequentist estimators of the *ω*_*n*_ values we have provided in Table 2 would be an excellent start towards this end, since these values stratified according to demographics could potentially be used in real time to optimise policies designed to mitigate the strength of non-adherence through, for example, targeted interventions.

## Data Availability

Data used were from the TUMIKIA project.

## Supporting information

**S1 Appendix**. Main mathematical content.

**S2 Appendix**. TUMIKIA project analysis and figures.

**S3 Appendix**. A comparison with existing models in the literature.

## Acknowledgements

The authors would like to sincerely thank Benjamin Collyer for careful reading of, and useful comments on, the manuscript. We extend our sincere thanks to the researchers who led data collection in Kenya during the TUMIKIA trial, including Paul M Gichuki, Stella Kepha, Carlos Mcharo, Stefan Witek-McManus, Mary W Karanja, Leah Musyoka, Lennie Mutisya, Tuva Safari, Idris Muye and Maureen Sidigu. Scientific oversight during data collection was provided by Charles Mwandawiro, Sammy Njenga and Simon Brooker. We also thank the many study participants who consented to provide stool samples, and the large study team, including managers, fieldworkers, laboratory technicians, and drivers. TUMIKIA was conducted in collaboration with the Government of Kenya Ministry of Health (MoH) and the Kwale County MoH, including Sultani Matendechero, Athuman Chiguzo, Hajara El-Busaidy and Redempta Muendo. Funding was received from the Bill & Melinda Gates Foundation, the Joint Global Health Trials Scheme of the Medical Research Council, the UK Department for International Development, the Wellcome Trust, and the Children’s Investment Fund Foundation. RJH, JET, MW and RMA gratefully thank the Bill & Melinda Gates Foundation for research grant support via the DeWorm3 (OPP1129535) award to the Natural History Museum in London (http://www.gatesfoundation.org/). The views, opinions, assumptions or any other information set out in this article are solely those of the authors. All authors acknowledge joint Centre funding from the UK Medical Research Council and Department for International Development.

## S1 Appendix

### Summary

In this supplementary information we derive the key mathematical expressions which are used and referred to in the main text.

#### Time-independent Markov model

Assuming that the conditional probabilities *β* and *γ* are constant, the time-independent Markov model may be mapped to the following recursion relation

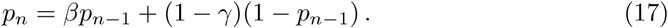

As in the main text, defining the system state vector as

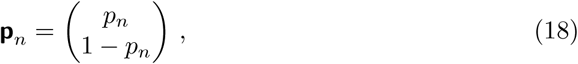

we may rewrite Eq (17) above in the form 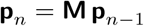 where we have defined the following transition matrix

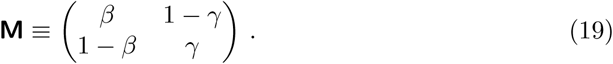

The eigenvalues and eigenvectors of **M** are given by

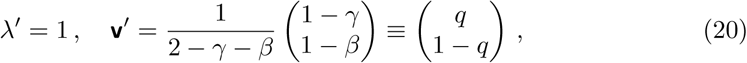

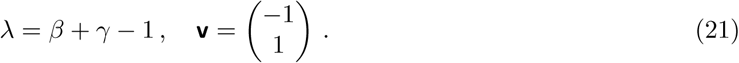

where **v**′ is normalised to sum to 1. Given that |*λ*| < 1 in all realistic circumstances, it is clear from this description that **v** represents the equilibrium of the system over multiple rounds with *λ* defining the rate of relaxation towards it. When *λ* = 0, the model becomes a history-independent model in which the next round is dictated solely by its probability at that round.

In order to study the dynamics in more detail, we apply the following transformation

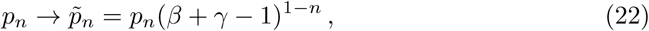

to the relation given by Eq (17), such that

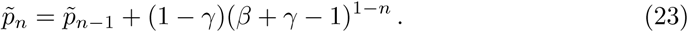

Through explicit summation, Eq (23) is solved by

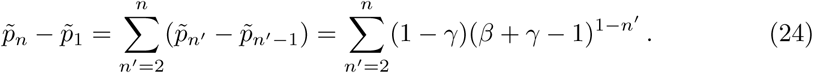

By reapplying the inverse transformation 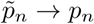 to Eq (24) and identifying 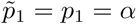, we obtain the following solution to Eq (17)

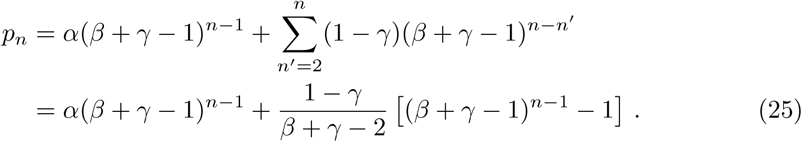

Equivalently, satisfying the dual to Eq (17) in terms of the probability of non-treatment in the *n*-th round 1 − *p*_*n*_, solutions to Eq (25) must also satisfy

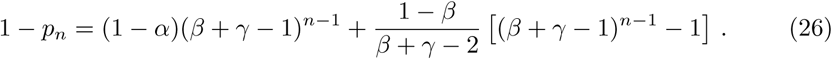

In Fig 3 we illustrate the dynamics of the system using Eq (25) with range of parameter values chosen for *γ*. Notice, in particular, that the system exhibits oscillation before relaxing to a steady state when *γ* is chosen such that the eigenvalue *λ* = *β* + *γ* − 1 < 0.

**Fig 3.**
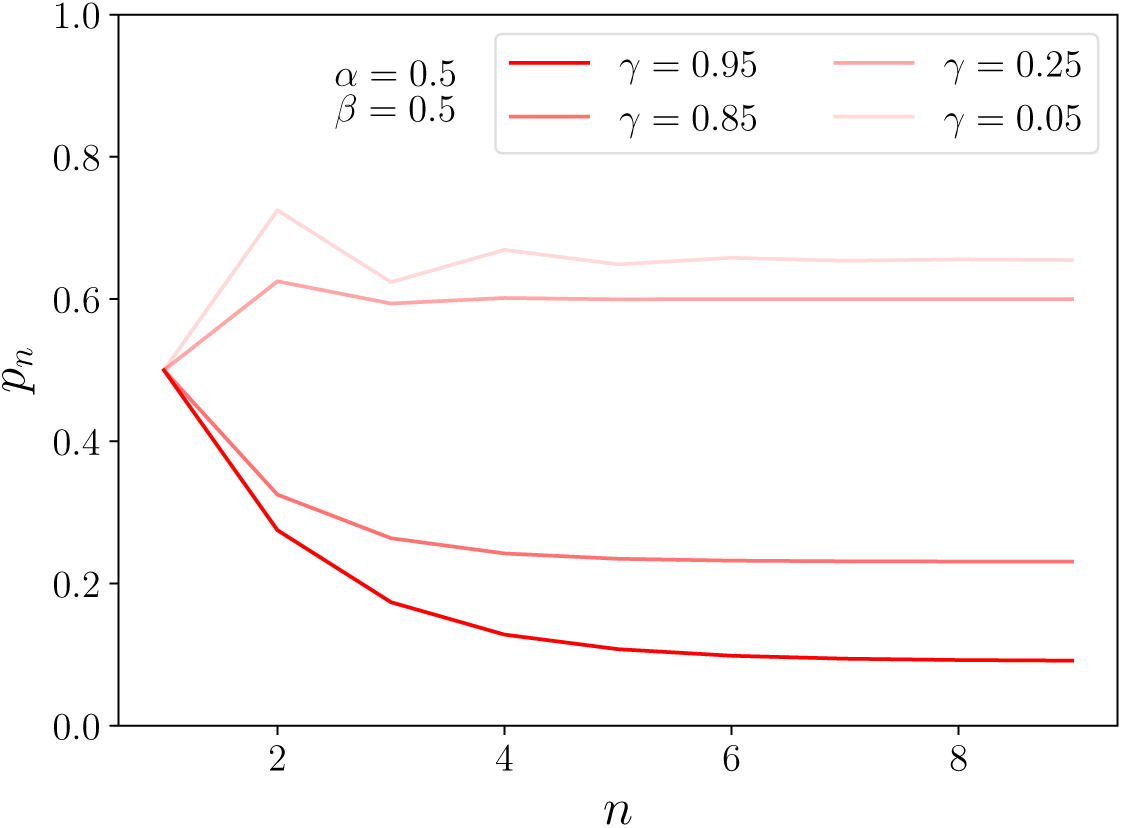
The probability of receiving treatment in the *n*-th round given by the Markovian model solution in Eq (25) for a range of *γ* values. The other probabilities have been fixed to *α* = 0.5 and *β* = 0.5.

For another way of calculating the expected lengths of repeat adherence E(*n*_T_) or non-adherence E(*n*_F_) of an individual (as computed in the main text), given that they begin with the same choice in the first round, one need only fix (*α* = *β, γ* = 1) or (*α* = 1 − *γ, β* = 1) and take moments with Eq (25), respectively, such that

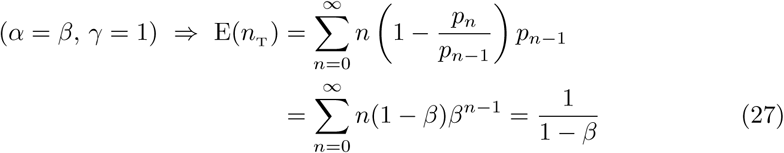

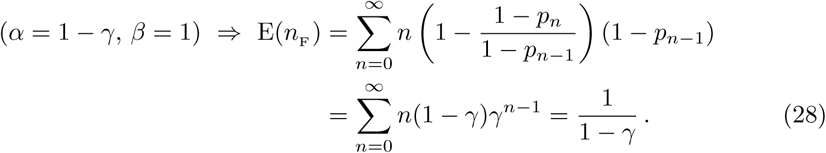

#### Time-dependent Markov model

Consider the choice matrices with elements 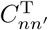 and 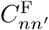 corresponding to the conditional probabilities of treatment and non-treatment in round *n* given treatment and non-treatment in round *n*′, respectively, such that

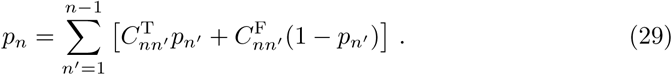

When the only nonzero elements of the choice matrices in Eq (29) are along the their lower diagonals, i.e., such that only 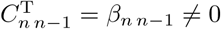 and 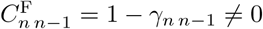 the system is described by a time-dependent Markov process with recursion relation

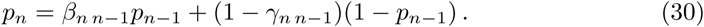

Following a similar argument to the one used in solving the homogeneous Markov case, we may obtain an implicit solution to Eq (30). Using the transformation

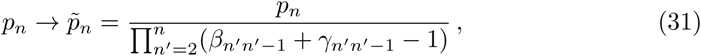

we once again substitute into the relation given by Eq (30), yielding

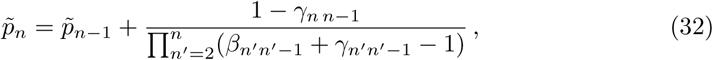

where Eq (32) is solved by the explicit summation

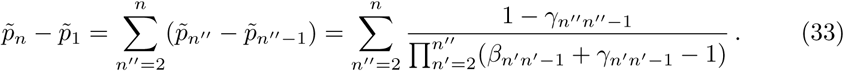

Using the corresponding inverse transformation to Eq (32) we hence obtain a solution to Eq (30), which is given by

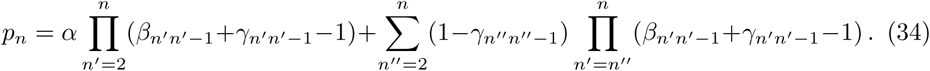

#### General choice matrices: non-Markovian models

The most general set of causal adherence models described by Eq (29) have choice matrices which take the form

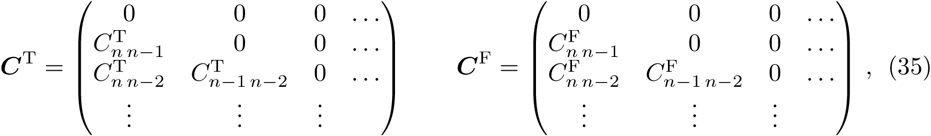

where ‘non-Markovian’ behaviour in the *n*-th round clearly corresponds to a past behaviour dependence between rounds which exceeds the immediate last round, i.e., 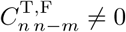 where *m* > 1.

Notice that all of the adherence models that we have identified in this work may be categorised by various constraints on the elements of the choice matrices introduced in Eq (29). For completeness and reference, these are

1. Past behaviour-independent adherence that is time-independent: ∀*n* > 1 only 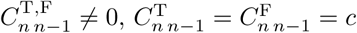 and *p*_1_ = *c*, giving one degree of freedom multiplied by the number of independent bins for population-level heterogeneity.
2. Past behaviour-independent adherence that is time-dependent: ∀*n* > 1 only 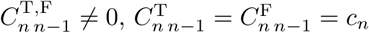 and *p*_1_ = *c*_1_, giving *n* degrees of freedom multiplied by the number of independent bins for population-level heterogeneity.
3. Markovian past behaviour-dependent adherence that is time-independent: ∀*n* > 1 only 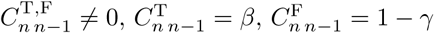and *p*_1_ = *α*, giving 2*n* − 1 degrees of freedom multiplied by the number of independent bins for population-level heterogeneity.
4. Markovian past behaviour-dependent adherence that is time-dependent: ∀*n* > 1 only 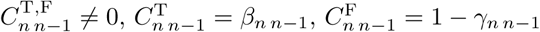 and *p*_1_ = *α*, giving 2*n* − 1 degrees of freedom multiplied by the number of independent bins for population-level heterogeneity.
5. Non-Markovian past behaviour-dependent adherence that is time-dependent: ∀*n* > 1 and ∀*n*′ < *n* only 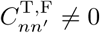 and *p*_1_ = *α*, giving 1 + *n*(*n* − 1) degrees of freedom multiplied by the number of independent bins for population-level heterogeneity.

#### Likelihoods and Bayesian evidence

Let the data now correspond to a set of *n*-vectors *D* = **{*X*}** where each individual’s adherence or non-adherence behaviour in the *n*-th round is recorded, such that *X*_*n*_ = T, F. Using Eq (29) the full generalisation of the likelihood (which supports all of the possible adherence models, becomes

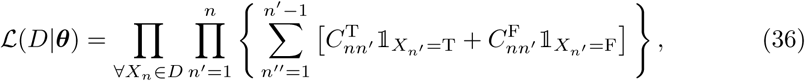

The large number of available degrees of freedom in Eq (36) motivates a systematic approach to inferring the choice matrix components from a given set of data. We elect to consider models which isolate the many degrees of freedom by constructing scenarios where past behaviour-dependent adherence only occurs for a single round and is temporally dependent on only one other round — all other degrees of freedom are hence set to those corresponding to time-dependent past behaviour-independent adherence, i.e. 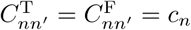. The likelihood for this more restricted set of models — which we denote as ℒ_*nn*_*′* (*D*|***θ***), where *nn*′ corresponds to the pair of rounds chosen to be dependent on each other in time — may be obtained by rewriting Eq (36) in the following form

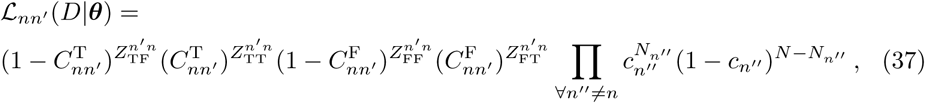

where we have defined

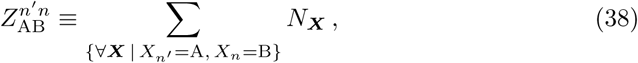

where the data *D* = {*N*_***X***_**}**has now been compressed into the set of numbers of people who track the same behaviour as ***X***, i.e., for 3 rounds, this forms the set of the following numbers of people: *N*_TTT_, *N*_TTF_, *N*_TFT_, etc. The Bayesian evidence integral corresponding to Eq (37) with a choice of flat prior *π*(***θ***) ∝ 1 is therefore

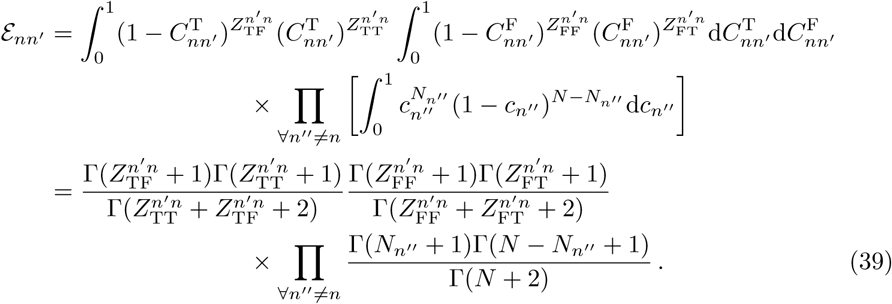

Some non-Markovian past dependence may be captured by the likelihood defined in Eq (37), however their Bayesian evidence may need to be compared with equivalent Markovian models which also generate decaying long-term correlations of a particular form. Using the same formalism as Eq (37), the time-dependent Markov model has the following likelihood

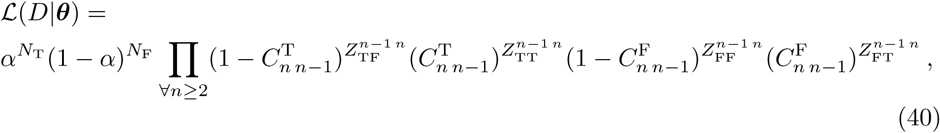

and, hence, yields the following Bayesian evidence

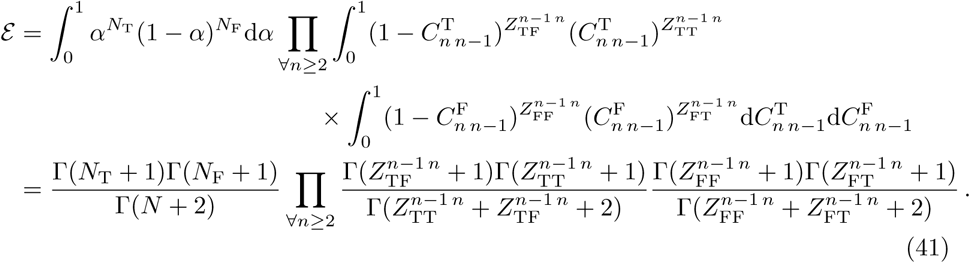

Eqs (40) and (41) may also be used to obtain the likelihood of the time-independent Markov model

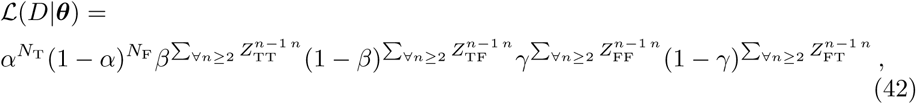

and the Bayesian evidence of the same model

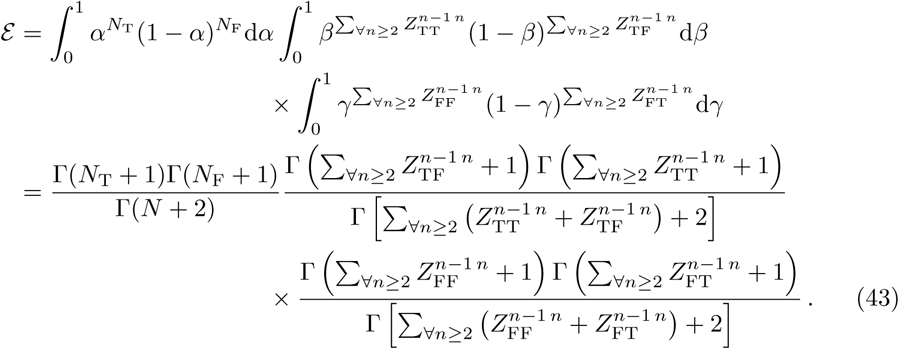

## S2 Appendix

### Summary

In this supplementary information we apply the framework of our mathematical model for adherence to the TUMIKIA project [11–13] and write a brief analysis description for each age group and sex.

### Introduction

In Figs 4, 5, 6, 7 and 8 we plot the maximum likelihood as well as the limits of the marginalised 95% credible region for the conditional probabilities given treatment (filled points) or non-treatment (hollow points) in a previous round of the overall, male and female participants in the top, middle and bottom rows, respectively. In the left column the constant conditional probabilities between any given sequential pair of rounds have been inferred, which corresponds to the time-independent Markov model of the main text and Appendix S1. In the right column all possible round pair dependencies are considered (indicated by the arrows on the horizontal axis), where in each case the components corresponding to a given round were measured assuming all other respective rounds were inferred to be from past behaviour-independent adherence. In all plots, above each pair of components we have also provided the log-Bayes factors [16], defined by

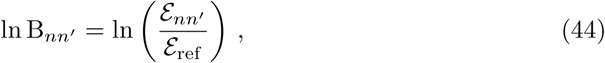

where the evidence for each pair *ε*_*nn*_*I* has been evaluated using the relations provided in Appendix S1 and the reference model evidence *ε*_ref_ has been set to that of time-dependent past behaviour-independent adherence for all components.

**Fig 4.**
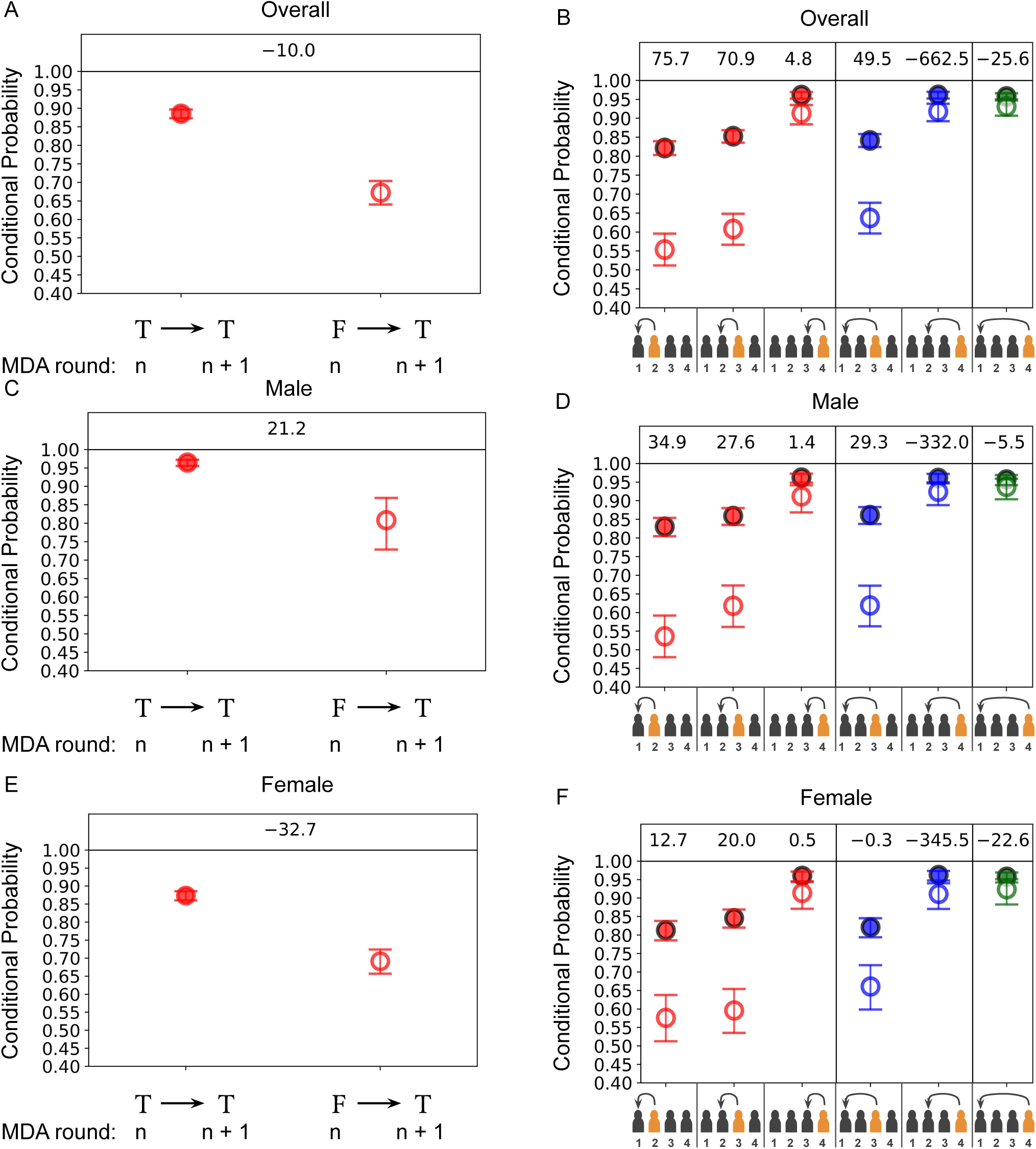
*Left column:* The maximum likelihood as well as the limits of the marginalised 95% credible region for the conditional probabilities of receiving treatment for any given pair of sequential rounds (these are hence homogeneous in time and the process is Markovian) given treatment (filled points) or non-treatment (hollow points) in a previous round. *Right column:* The same as the left column but with allowed time dependence in the conditional probabilities of receiving treatment in each respective round (highlighted in orange on the horizontal axes). In each case the components corresponding to a given round were measured assuming all other respective rounds were inferred to be from time-dependent past behaviour-independent adherence and hence the likelihood is given in Appendix S1. Different colours for each point correspond to different lengths in time for the dependencies in behaviour. The datasets used are from the standard pre-SAC (0-4) age category from a cohort of individuals from the biannual treatment arm of the TUMIKIA project where the: top row corresponds to the overall group; middle row corresponds to the male sub-group; and bottom row corresponds to the female sub-group.

**Fig 5.**
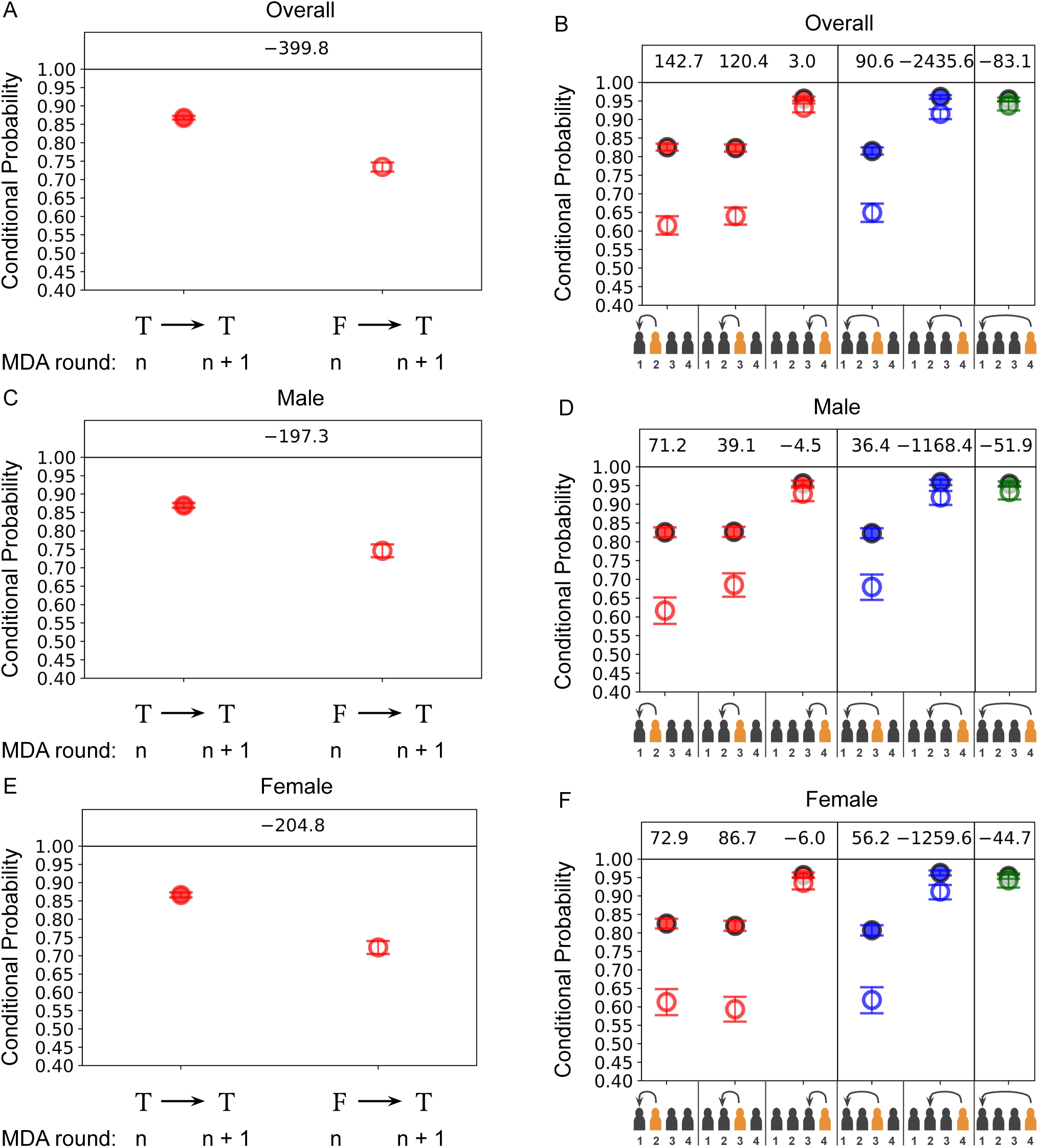
*Left column:* The maximum likelihood as well as the limits of the marginalised 95% credible region for the conditional probabilities of receiving treatment for any given pair of sequential rounds (these are hence homogeneous in time and the process is Markovian) given treatment (filled points) or non-treatment (hollow points) in a previous round. *Right column:* The same as the left column but with allowed time-dependent in the conditional probabilities of receiving treatment in each respective round (highlighted in orange on the horizontal axes). In each case the components corresponding to a given round were measured assuming all other respective rounds were inferred to be from time-dependent past behaviour-independent adherence and hence the likelihood is given in Appendix S1. Different colours for each point correspond to different lengths in time for the dependencies in behaviour. The datasets used are from the standard SAC (4-15) age category from a cohort of individuals from the biannual treatment arm of the TUMIKIA project where the: top row corresponds to the overall group; middle row corresponds to the male sub-group; and bottom row corresponds to the female sub-group.

**Fig 6.**
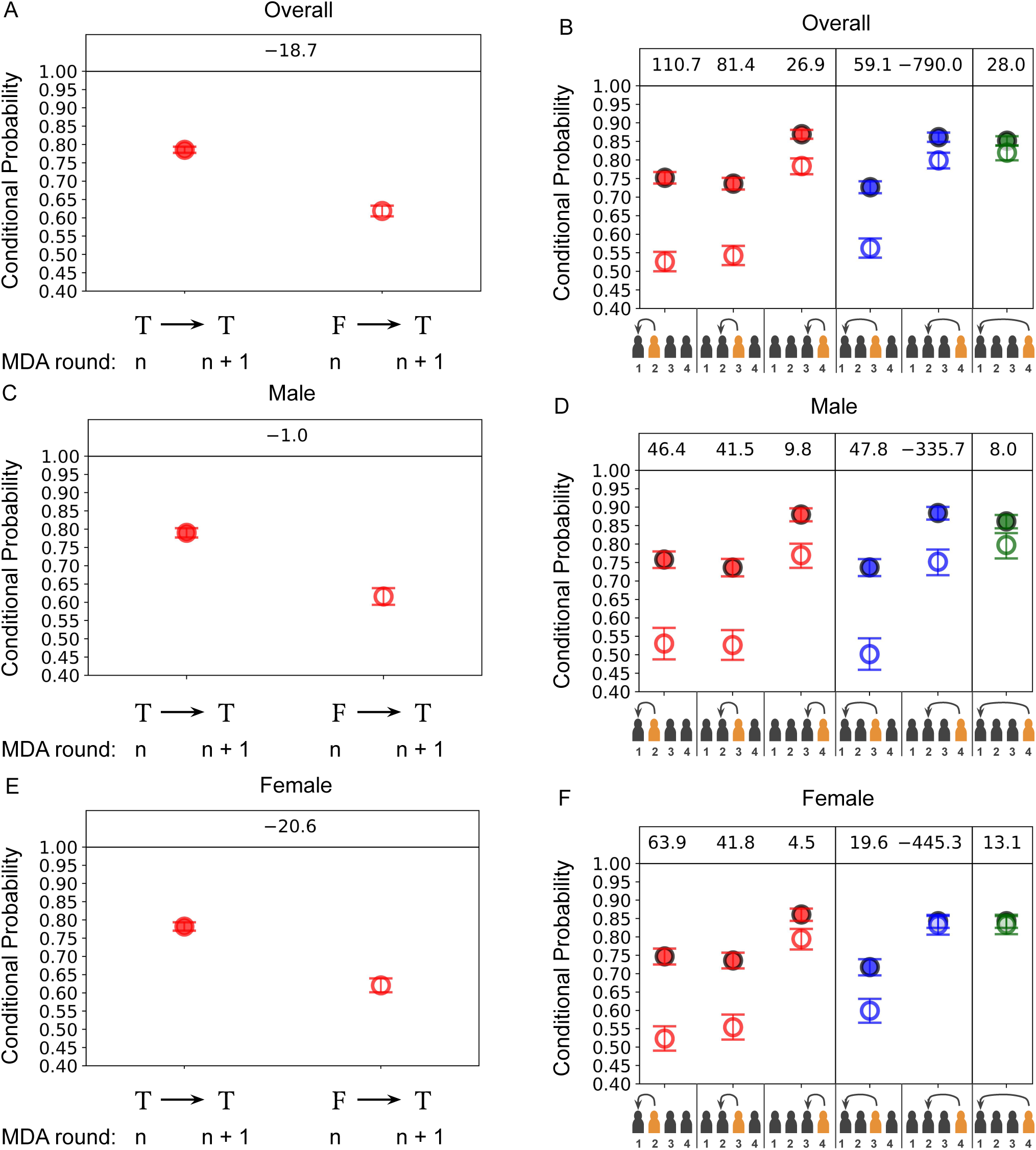
*Left column:* The maximum likelihood as well as the limits of the marginalised 95% credible region for the conditional probabilities of receiving treatment for any given pair of sequential rounds (these are hence homogeneous in time and the process is Markovian) given treatment (filled points) or non-treatment (hollow points) in a previous round. *Right column:* The same as the left column but with allowed time-dependent in the conditional probabilities of receiving treatment in each respective round (highlighted in orange on the horizontal axes). In each case the components corresponding to a given round were measured assuming all other respective rounds were inferred to be from time-dependent past behaviour-independent adherence and hence the likelihood is given in Appendix S1. Different colours for each point correspond to different lengths in time for the dependencies in behaviour. The datasets used are from the 15-29 age category from a cohort of individuals from the biannual treatment arm of the TUMIKIA project where the: top row corresponds to the overall group; middle row corresponds to the male sub-group; and bottom row corresponds to the female sub-group.

**Fig 7.**
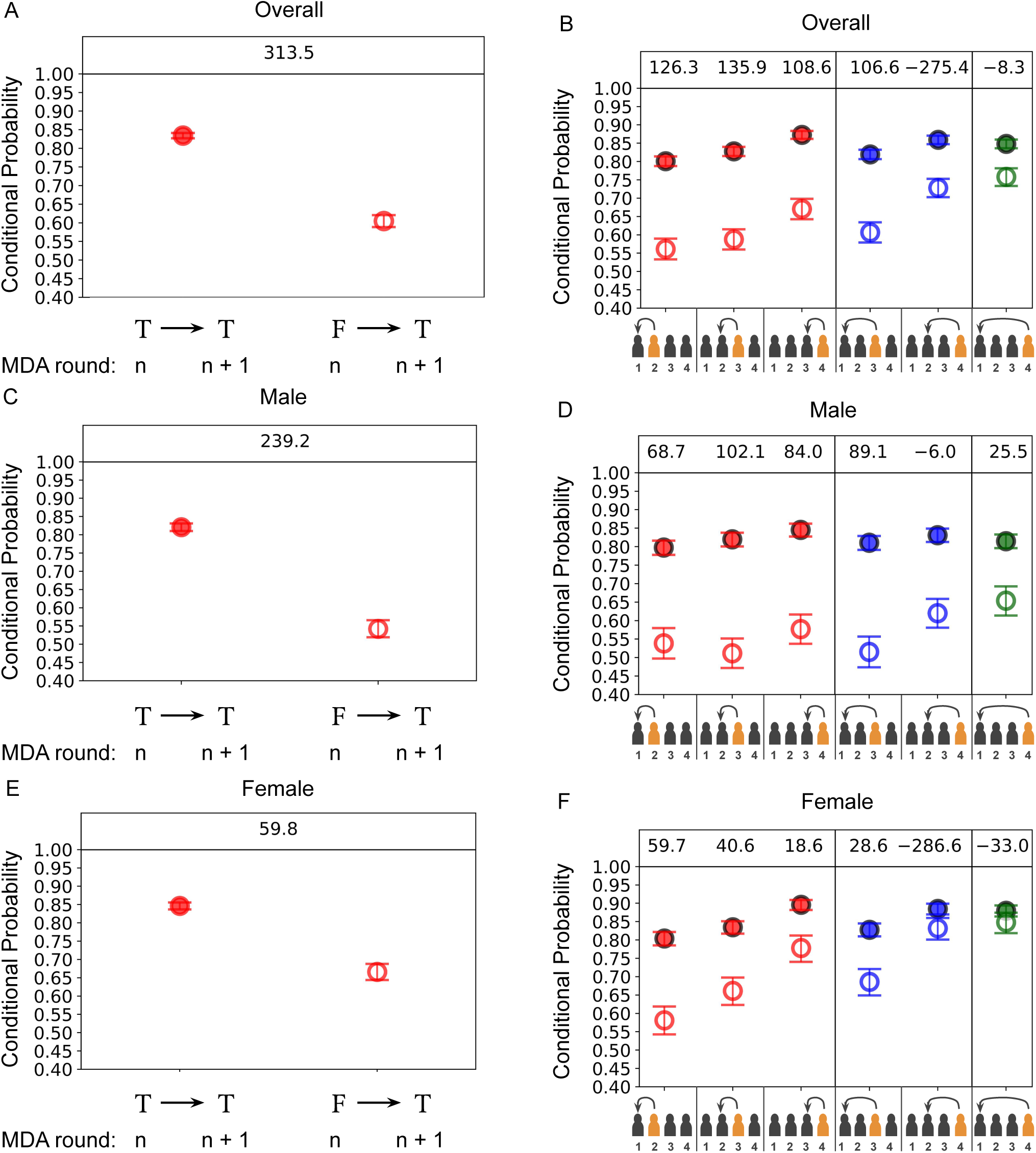
*Left column:* The maximum likelihood as well as the limits of the marginalised 95% credible region for the conditional probabilities of receiving treatment for any given pair of sequential rounds (these are hence homogeneous in time and the process is Markovian) given treatment (filled points) or non-treatment (hollow points) in a previous round. *Right column:* The same as the left column but with allowed time dependence in the conditional probabilities of receiving treatment in each respective round (highlighted in orange on the horizontal axes). In each case the components corresponding to a given round were measured assuming all other respective rounds were inferred to be from time-dependent past behaviour-independent adherence and hence the likelihood is given in Appendix S1. Different colours for each point correspond to different lengths in time for the dependencies in behaviour. The datasets used are from the 30-49 age category from a cohort of individuals from the biannual treatment arm of the TUMIKIA project where the: top row corresponds to the overall group; middle row corresponds to the male sub-group; and bottom row corresponds to the female sub-group.

**Fig 8.**
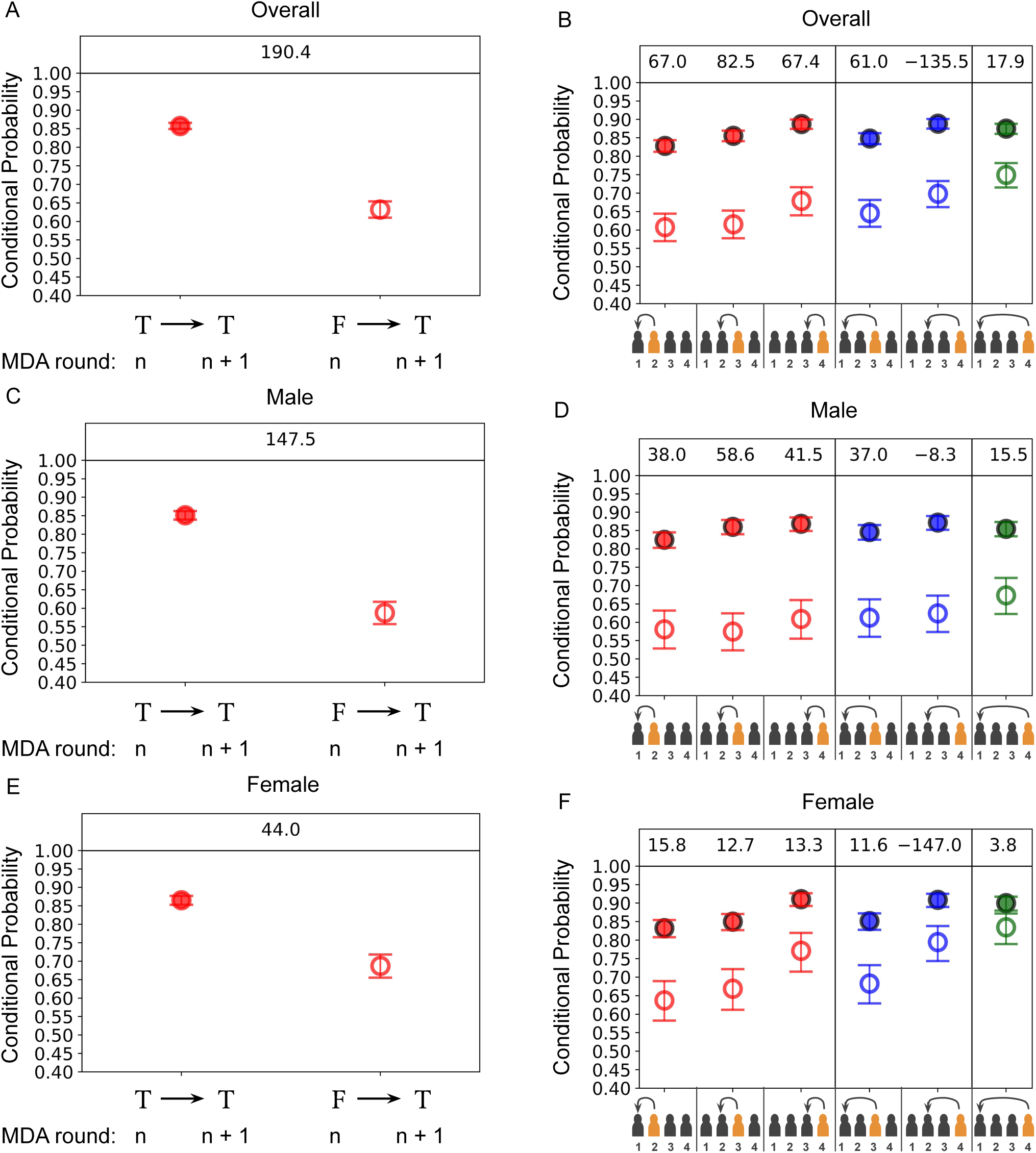
*Left column:* The maximum likelihood as well as the limits of the marginalised 95% credible region for the conditional probabilities of receiving treatment for any given pair of sequential rounds (these are hence homogeneous in time and the process is Markovian) given treatment (filled points) or non-treatment (hollow points) in a previous round. *Right column:* The same as the left column but with allowed time dependence in the conditional probabilities of receiving treatment in each respective round (highlighted in orange on the horizontal axes). In each case the components corresponding to a given round were measured assuming all other respective rounds were inferred to be from time-dependent past behaviour-independent adherence and hence the likelihood is given in Appendix S1. Different colours for each point correspond to different lengths in time for the dependencies in behaviour. The datasets used are from the 50+ age category from a cohort of individuals from the biannual treatment arm of the TUMIKIA project where the: top row corresponds to the overall group; middle row corresponds to the male sub-group; and bottom row corresponds to the female sub-group.

### Results

In Figs 4, 5 and 6 we present our results for the pre-SAC, SAC and 15-29 age groups of individuals in the TUMIKIA project. These age groups appear to be well-described by a time-dependent Markov model so past behaviour-dependent non-adherence is clearly present. This may be identified by the largest log-Bayes factor values being given in the red-coloured right column plots for all three sets of plots. However, the conditional probabilities in all groups appear to drift closer together by round 4 of treatment, which signals a gradual transition from past behaviour-dependent to independent adherence.

In Figs 7 and 8 we present our results for the 30-49 and 50+ age groups of individuals in the TUMIKIA project. The overall cohort, as well as the males and females in both age groups, appear to exhibit strong evidence of past behaviour-dependent non-adherence — in particular, they are all apparently well-described by a time-independent Markov model. These conclusions may be drawn both by the consistent distance between all of the values for the inferred conditional probabilities with the red points of the right column of plots, as well as the largest evidence (as measured by the log-Bayes factor in the top row of the plots) for a difference in conditional probabilities in the left column in both plots.

In all of the cohorts studied in Figs 4, 5, 6, 7 and 8, we report no evidence for the existence of dependencies between rounds that depart from a Markovian description (as can be inferred from the comparatively small log-Bayes factors for the blue and green conditional probabilities in the right column of all plots). This is an interesting, and perhaps surprising, result regarding the nature of human behaviour in response to mass drug administration.

## S3 Appendix

### Summary

In this supplementary information, we analyse some of the existing models of adherence from the literature in the context of our proposed framework.

#### The Plaisier model

Several models of MDA treatment programmes employ an adherence model developed by Plaisier in the context of onchocerciasis control [8, 20]. The Plaisier model assigns a propensity score for adherence to each individual which they then retain for the duration of the MDA programme [14, 15]. In the time-independent form, this model would be characterised by us as a heterogeneous population, time-independent model with no explicit individual dependence on past behaviour — though more recently developed models include time dependent effects through coverage variability between rounds. In the simplest model, the individual probability of adherence is given by *U* ^(1−*c*)*/c*^, where *U* is a uniform random number and *c* is expected probability of treatment and hence the expected coverage. The model is therefore completely parameterized by the overall expected coverage. The PDF for the adherence probability for this process is given by

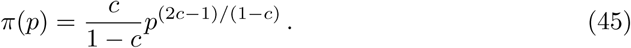

The PDF of *p* rises monotonically from zero to one for all values of *c* > 0.5 and falls monotonically for *c* < 0.5 (for *c* = 0.5, it is flat). Note that *π*(*p*) is a beta distribution: *π*(*p*) = Beta[*p*; *c/*(1 − *c*), 1]. For this distribution, the mean failure run length is hence given by

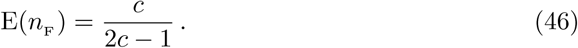

Note that in this model, adherence failure run length becomes undefined at a coverage of 50% or less. Additionally, one can show that the variance of this random variable becomes undefined for values of coverage below 66%, suggesting that failure run lengths in finite populations drawn from this distribution will exhibit extreme variability.

The probability of an individual being untreated across *N* rounds of MDA in this model can also be calculated, giving

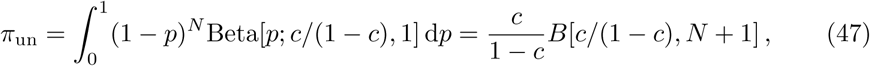

where *B*(,) is the beta function. Fig 9 shows the distribution of adherence probabilities for 2 different coverage values and also the probability of an individual not adhering with treatment across a 4-round MDA programme.

**Fig 9.**
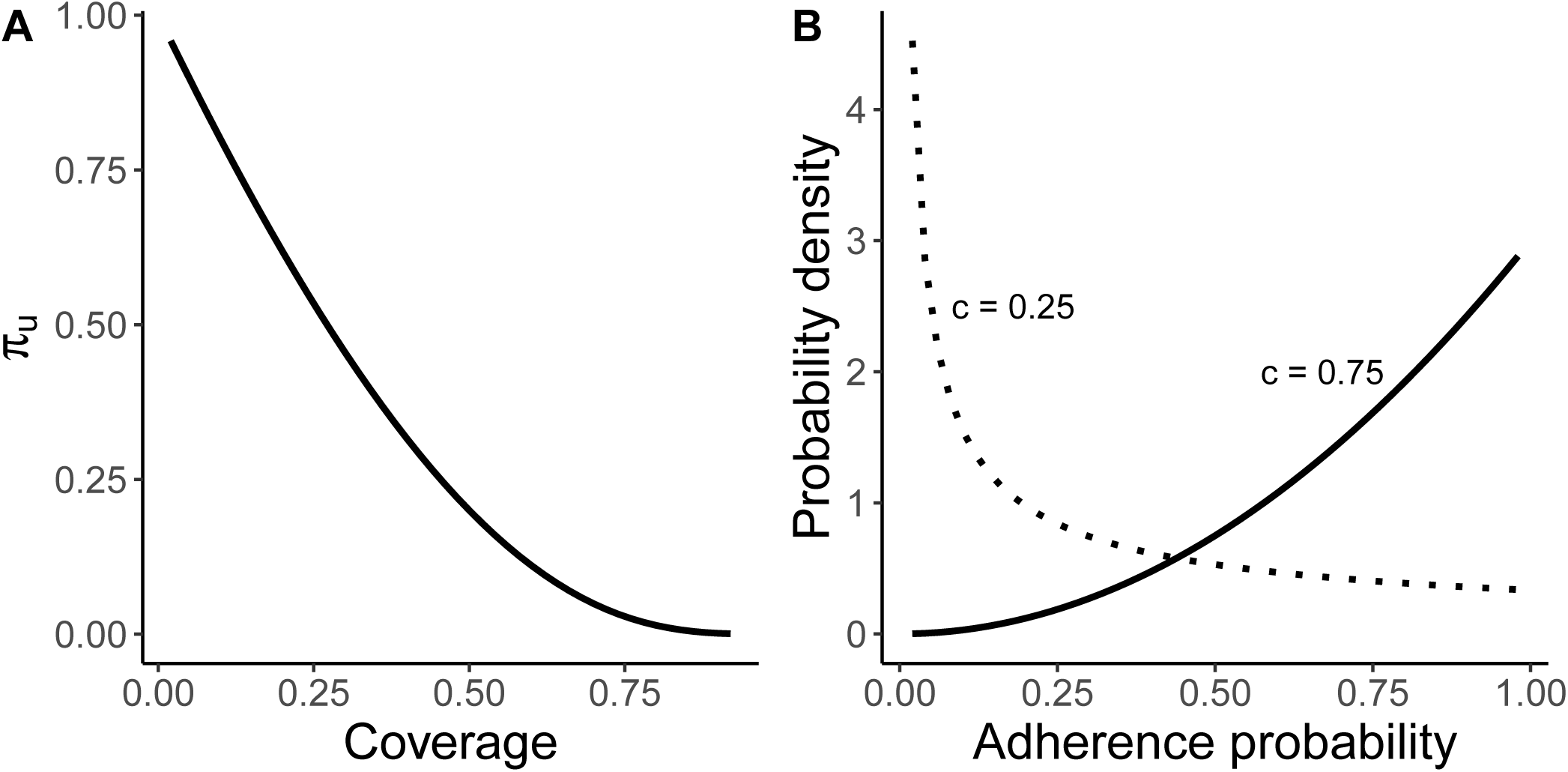
A) Probability of an individual with adherence drawn from the Plaisier distribution of not adhering with treatment during a 4 round MDA programme. B) The probability distribution for adherence for coverages of 25% and 75%.

Note here that in order to fully implement the Plaiser model in our framework, the additional variability in time through coverage would have to be added to our description. This should be straightforward to do within our proposed formalism.

#### The Griffin Model

The adherence model used by Irvine et al [5] to model MDA adherence in the treatment of lymphatic filariasis was originally created by Griffin et al in the context of intervention strategies against malaria transmission [21]. The original Griffin model is quite broad and deals with multiple simultaneous interventions and the correlations in their uptake. It does not include conditional dependencies for an individual’s behaviour and is therefore a heterogeneous population, time-independent, individually past behaviour-independent model in its simplest form. Each individual in the population is assigned a correlation parameter, *u*_*i*_, drawn from a normal distribution with mean *u*_0_ and variance *σ*^2^. These parameters are retained throughout the MDA programme. At each round a MDA round, each individual draws a unit-variance normal deviate with mean *u*_*i*_, *z*. Treatment is received if *z* < 0. The expected coverage is given by 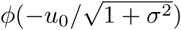, where *ϕ* is the standard normal cumulative probability function. This leaves one free parameter to control the distribution of adherence probabilities across the population.

The cumulative distribution of adherence probability, *p*, is given by

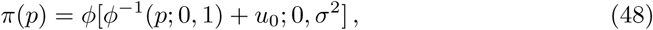

giving a PDF

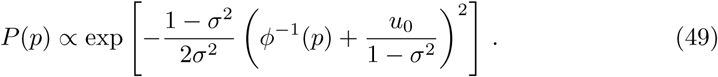

The function *ϕ*^−1^(*p*; 0, 1) varies monotonically in the range (− ∞, ∞) with *p*. In Eq (49), the parameter *σ* = 1 acts to discriminate between two functional forms. For *σ* < 1, the distribution has a ‘normal’ shape with a single local maximum, while for *σ* > 1, the distribution has asymptotes with local maxima at the *p* = 0 and/or 1. In this, it is very similar, qualitatively, to the beta distribution (see Fig 10).

**Fig 10.**
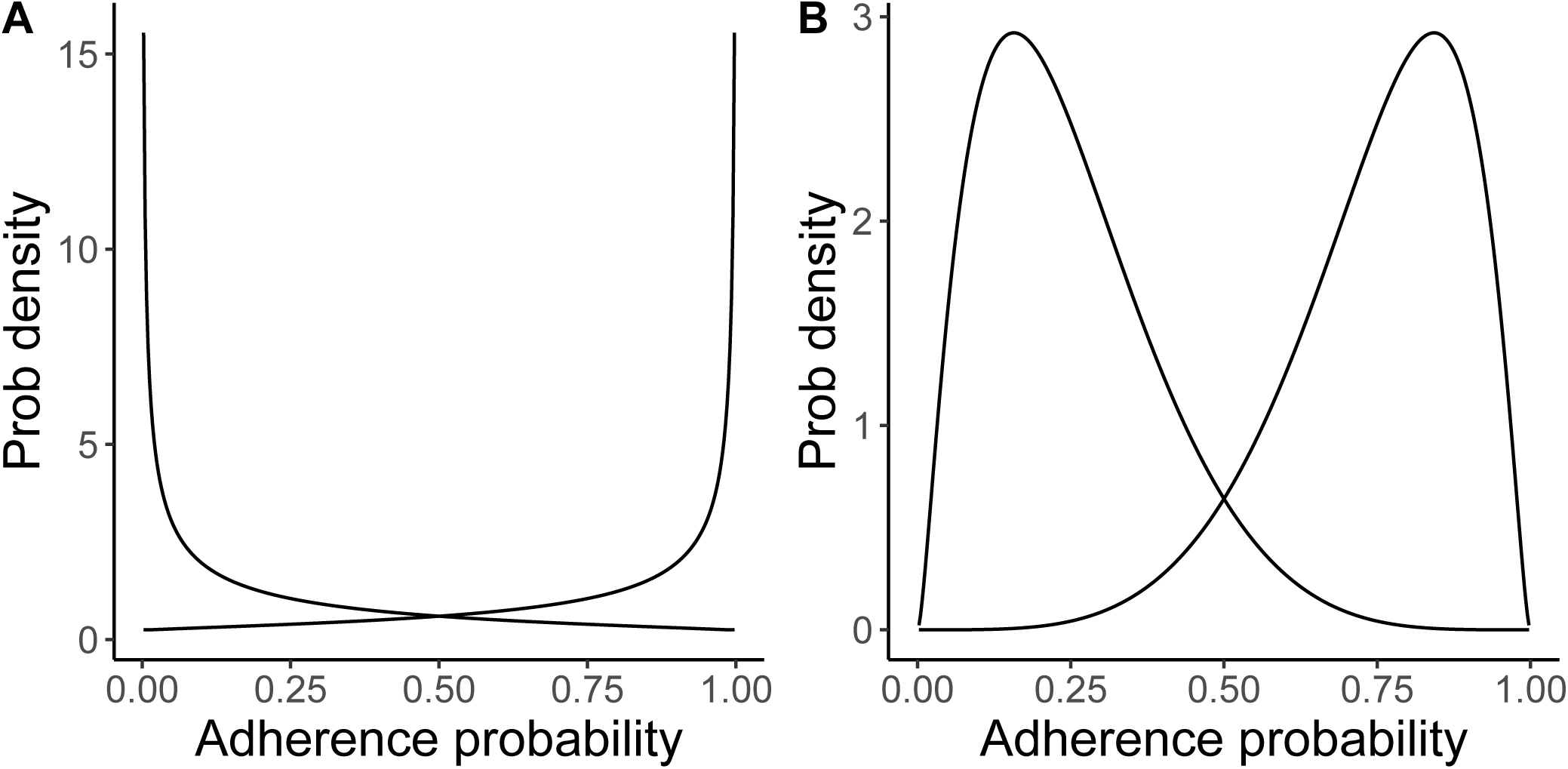
Adherence probability distributions with A) *σ* = 1.2 and B) *σ* = 0.5 for mean coverages of 25% and 75%. The probability distribution for adherence for coverages of 25% and 75%.

